# Encoding of pretrained large language models mirrors the genetic architectures of human psychological traits

**DOI:** 10.1101/2025.03.27.25324744

**Authors:** Bohan Xu, Nick Obradovich, Wenjie Zheng, Robert Loughnan, Lucy Shao, Masaya Misaki, Wesley K. Thompson, Martin Paulus, Chun Chieh Fan

## Abstract

Recent advances in large language models (LLMs) have prompted a frenzy in utilizing them as universal translators for biomedical terms. However, the black box nature of LLMs has forced researchers to rely on artificially designed benchmarks without understanding what exactly LLMs encode. We demonstrate that pretrained LLMs can already explain up to 51% of the genetic correlation between items from a psychometrically-validated neuroticism questionnaire, without any fine-tuning. For psychiatric diagnoses, we found disorder names aligned better with genetic relationships than diagnostic descriptions. Our results indicate the pretrained LLMs have encodings mirroring genetic architectures. These findings highlight LLMs’ potential for validating phenotypes, refining taxonomies, and integrating textual and genetic data in mental health research.

## Introduction

Large language models (LLMs) have demonstrated remarkable advancements in recent years ^1^, fueling enthusiasm for their potential as versatile tools across a variety of domains, including biomedical and clinical research. These models, trained on vast corpora of text, have been proposed for tasks ranging from extracting and linking information from electronic medical records to generating embeddings that predict clinical outcomes ^2,3^. Yet, fundamental questions remain: What do LLMs truly encode within their latent spaces? And how can we rigorously benchmark these representations at scale?

To explore these questions, we examined whether LLMs can implicitly capture biologically grounded relationships among psychological and psychiatric constructs. Specifically, we focused on the “zero-shot” performance of LLM embeddings—how well they reflect relationships between concepts without fine-tuning. To provide a biologically informed benchmark, we leveraged population-level genetic correlations (GCs) ^4^, which quantify the shared effects of single nucleotide polymorphisms (SNPs) between traits or disorders. GCs offer a robust measure of biological relatedness, making them an ideal reference for evaluating whether LLM embeddings capture meaningful patterns ^5,6^.

We specifically probed: *To which degree, without additional training, the LLM-based textual embeddings of items from a neuroticism questionnaire and DSM-5 diagnostic categories reflect their genetically defined relationships?* The convergence of textual embeddings with GCs would indicate that LLMs are not merely encoding arbitrary or idiosyncratic semantic associations but instead are aligning with human biology. In contrast to the frequently used text retrieval benchmarks ^7^, which evaluate the accuracies of the text queries given manually designed artificial datasets, our results demonstrate the potential to bridge two disparate knowledge domains—textual corpora and genetic data—shedding light on how statistical patterns in language reflect, and sometimes mirror, the biological organization of traits and disorders.

## Results

### Examining the alignment between text embedding and biological embedding for Neuroticism questionnaire items

We selected 10 different LLM text embedding models, eight of them being state-of-the-art encoder models (as of this writing) available in recent AI booms, with the other two being popular open-source models (detailed in **Supplementary Table 1**). Detailed procedures of the text embedding, as well as the comparisons with the estimated GCs can be found in the **Online Methods**. **Figure 1a** illustrates the procedural steps we used to compare the text embeddings from 10 different LLM models with the biological embeddings based on genetic correlations estimated using a population cohort. The resulting performance metric, i.e. how much of the variation in GCs between 12 items of a neuroticism questionnaire (**Supplementary Table 2**) ^8^ can be explained by zero-shot text embeddings of those questionnaire items, is shown in the **Figure 1b** and **Figure 1c**.

**Figure 1.**
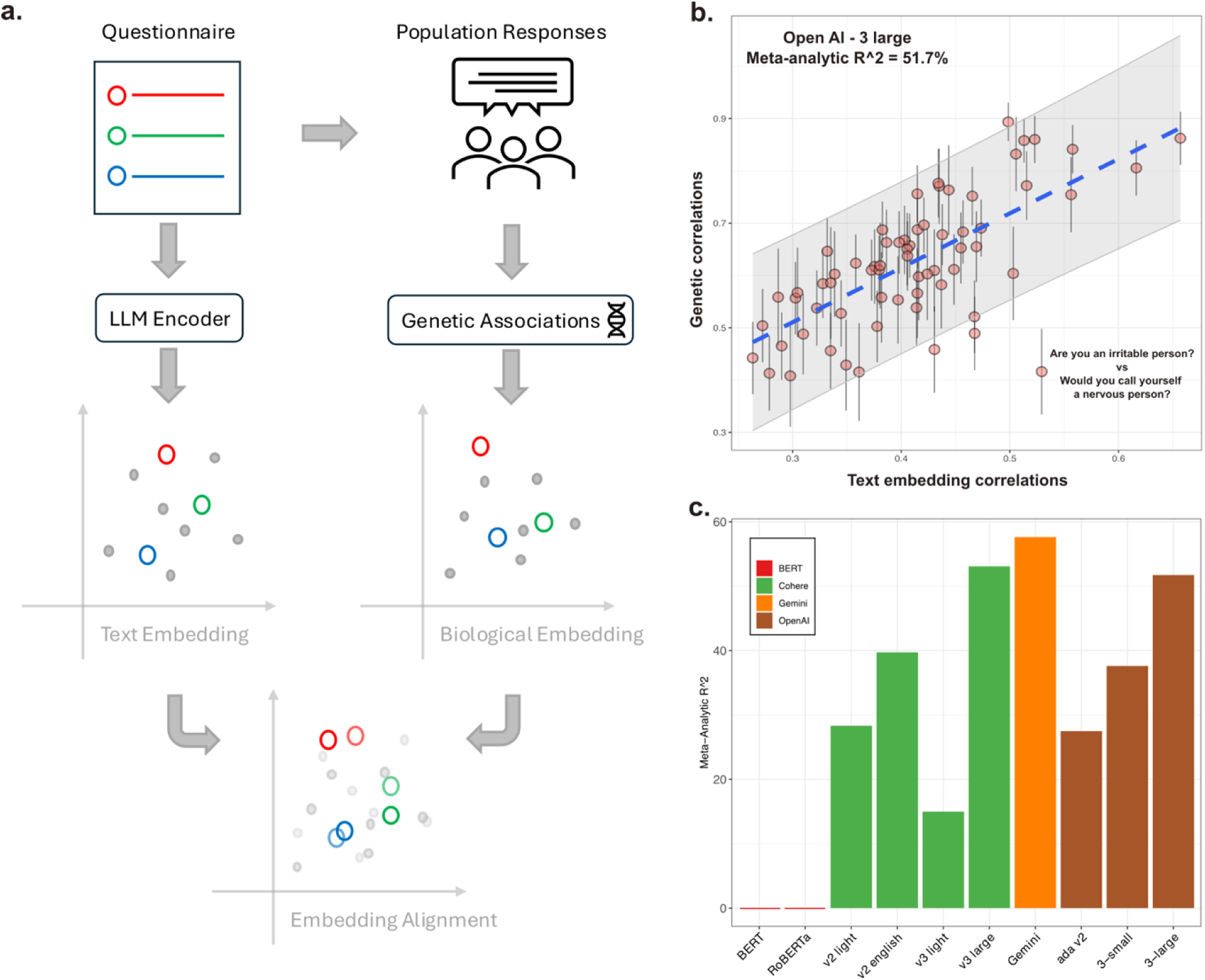
Comparisons between text embedding and genetically defined biological embedding. a). Conceptual framework of current study. b). Example of the alignment between text embedding and genetic correlations. The text embedding was derived using OpenAI 3-large model. Each point represents one pair of the questionnaire items. 95% confidence intervals of the genetic correlation estimates were provided. c). Variance of genetic correlations explained by zero-shot text embedding from the LLMs, estimated with linear mixed effects meta-regression. Colored by the model groups.

Figure 1b showcases a detailed view of the text embeddings derived from the OpenAI’s v3 large-dimension model, demonstrating remarkable alignment with the empirical GCs of neuroticism items estimated from large-scale population cohort ^8,9^. Notably, the embeddings captured the clustering of specific items that share stronger heritable relationships, such as “mood swings” and “nervousness,” which exhibited high genetic correlation in the UK Biobank data. The robustness of this alignment is underscored by the absence of major outliers, even when meta-regressions accounting for uncertainty in the genetic estimates (Figure 1b). The neuroticism questionnaire’s well-validated psychometric properties ^8^ and high-powered genetic estimates (N ≈ 84,000–100,000 for item-level GWAS) likely contributed to the strong alignment observed. This suggests that the semantic relationships encoded by LLMs reflect the latent structure of how individuals respond to these items in a biologically meaningful way.

Across 10 selected LLMs, larger models, particularly those without dimensionality reduction, demonstrated a clear performance advantage over smaller models (Figure 1c). For instance, OpenAI’s v3 large-dimension model captured 51% of the variance of the genetic embeddings, compared to 37% for the dimensionally reduced v3 model and 27% for the older ada v2 model (Figure 1c). This progressive improvement highlights the importance of model scale and dimensionality in encoding biologically grounded relationships.

### Psychiatric Disorders: Disorder Names vs. DSM-5 Criteria

When examining the six major psychiatric disorders (Major Depressive Disorder, Generalized Anxiety Disorder, Bipolar disorder, Schizophrenia, Attention Deficit and Hyperactivity Disorder, and Post-Traumatic Stress Disorder), we found that the text embeddings of disorder names captured genetic relationships more effectively than embeddings based on the descriptive criteria A of DSM-V (Figure 2a and **2b**). Embedding of the DSM-5 descriptions showed miniscule alignment (Figure 2a), whereas the text embedding based on the diagnostic name only capture up to 29% of the variance of the GCs (Figure 2b). The superior performance of disorder names suggests that the textual data from which LLMs derive their embeddings may better capture co-occurrence patterns and conceptual associations in real-world contexts than does the semantic information from the diagnostic descriptions. This observation is consistent with the idea that real-world discourse more accurately reflects underlying biological relationships, perhaps because it implicitly incorporates relationships between comorbidities and shared etiological pathways.

**Figure 2.**
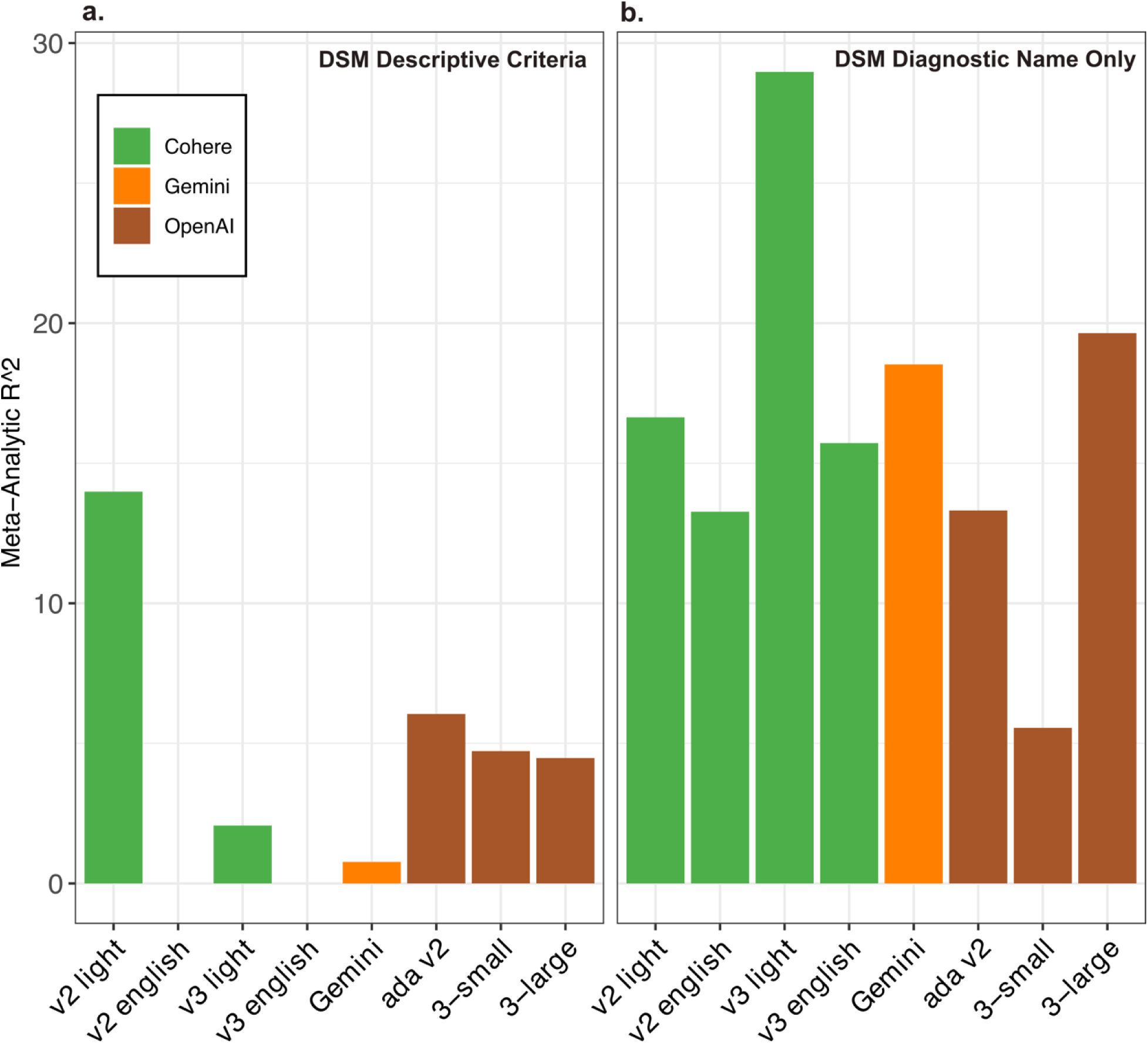
Variance of genetic correlations between psychiatric disorders explained by zero-shot text embedding from LLMs. a). Results based on encoding the descriptions from DSM-V diagnostic criteria. b). Results based on encoding the diagnostic names from DSM-V, e.g. schizophrenia and major depressive disorder.

## Discussion

Our findings demonstrate that pretrained encoder-only large language models (LLMs) can approximate biologically grounded relationships among psychological traits and psychiatric disorders with surprising fidelity, even in a zero-shot setting. These models, trained solely on vast text corpora, encode emergent properties in their semantic spaces that reflect population-level patterns observed in genetic correlations (GCs). For example, embeddings from the largest OpenAI model tested captured 51% of the variance in the GC structure for neuroticism items, suggesting that LLMs are capable of encoding meaningful relationships rooted in human biology.

This study highlights the potential of population genetic data to serve as a novel benchmark for evaluating LLM embeddings. Unlike traditional benchmarks based on human annotations or curated tasks, genetic correlations provide large-scale, biologically grounded indices that are relatively unbiased by selection processes. The alignment between LLM embeddings and GCs suggests that these models can uncover latent structures in language that mirror real-world, biologically meaningful relationships. Such benchmarks could enable systematic evaluations of how well LLMs represent relationships across domains, moving beyond tasks focused solely on linguistic accuracy or surface-level semantics.

A key observation is that LLMs more effectively capture genetic relationships when encoding disorder names than when using DSM-5 diagnostic criteria. This finding suggests that the statistical patterns in real-world language use—across scientific articles, medical forums, and general texts— reflect an implicit taxonomy of psychological and psychiatric constructs. This taxonomy, shaped by how humans naturally discuss and group concepts, may already encode partial knowledge of their genetic and neurobiological relationships. In contrast, the weaker alignment of DSM-5 criteria highlights potential limitations in formal diagnostic systems. These criteria are linguistically complex, often overlapping, and rooted in consensus rather than strictly biological underpinnings. The discrepancy underscores the importance of reconsidering how psychiatric knowledge is formalized and communicated, particularly in ways that might better align with emerging biological insights.

These findings mark a significant step toward integrating textual data with biological insights to advance mental health research. The convergence of LLM embeddings with population-level genetic correlations reveals the potential of AI systems to uncover meaningful relationships in human health and behavior. This cross-disciplinary approach holds promise for refining psychiatric taxonomies, developing personalized medicine tools, and inspiring new methodologies for questionnaire design and phenotypic discovery. Moving forward, the ability of LLMs to highlight undiscovered or underexplored biological relationships offers a unique opportunity for hypothesis generation and refinement of diagnostic frameworks. By systematically identifying trait or disorder pairs with high semantic similarity but little-known genetic overlap, researchers could conduct targeted genome-wide association studies to confirm potential shared heritability. Such zero-shot insights may also spur the development of more biologically informed psychiatric taxonomies that align with how disorders are discussed in natural language, moving beyond formal diagnostic criteria that often struggle to capture genetic and neurobiological subtleties. Iterative validation in prospective clinical studies—integrating patient risk profiles, real-world discourse, and genetic data—could then help validate these newly discovered relationships, ensuring that any resultant shifts in nosology or treatment guidelines are both biologically grounded and clinically applicable.

## Data Availability

All data produced in the present work are contained in the manuscript

## Online Methods

### Items used in the embedding queries

We selected a Neuroticism questionnaire as our main text embedding query due to their psychometric properties and readily available large-scale genome-wide association results ^8,10^. Genetic correlations are likely more accurate given a well powered study design. The neuroticism questionnaire was made up of 12 items from the Eysenck Personality Questionnaire, Revised Short Form (EPQ-RS) ^11^ - the exact text of the questionnaire items used in this study can be found in **Supplementary Table 1**. Six psychiatric disorders which have genetic correlations estimated in the same genome-wide association study ^8,10^ are included in our analyses, which are Major depressive disorder, Schizophrenia, Anorexia Nervosa, Autism, Bipolar I Disorder, and Attention-Deficit/Hyperactivity Disorder. We derived their text embedding based on either their descriptive criteria from Diagnostic and Statistical Manual of Mental Disorders, Fifth Edition, Text Revision or their diagnostic name only. Detailed text fed into the LLMs can also be found in **Supplementary Table 1**.

### Encoder-only Large Language Models

Unlike encoder-decoder and decoder-only models designed for generative tasks like ChatGPT, embedding models (encoders) focus on transforming input sentence, into a meaningful numerical representation known as an embedding. While all these models utilize the attention mechanism ^12^, their primary differences are the architecture and pretraining tasks ^13–16^. Decoders employ masked attention, restricting access to only earlier positions in the output sequence, which enables sequential generation. In contrast, encoders process the entire input sequence simultaneously, capturing holistic contextual information. Decoders are typically pretrained using Causal Language Modeling (CLM), which involves predicting the next token (the fundamental unit in LLMs) in a sequence based on prior tokens ^14,15^. In contrast, encoders commonly use Masked Language Modeling (MLM) for pretraining, where tokens in the input sequence are randomly masked, and the model is trained to predict these masked tokens ^13,16^. In the following sections, we described the encoder-only models we applied in this study.

#### Open-source models

Bidirectional Encoder Representations from Transformers (BERT) ^13^ is the first model to leverage the self-attention mechanism while jointly conditioning on both left and right context, for creating vector representations of language. The initial release of BERT includes two variants, where BERT_BASE_ has about 110M parameters while BERT_LARGE_ has approximately 340M parameters. Both models are pre-trained with same data, BooksCorpus and English Wikipedia, as well as same tasks, MLM and Next Sentence Prediction (NSP).

Liu et al. ^16^ introduced RoBERTa (Robustly Optimized BERT Approach), a pretraining and optimization strategy designed to enhance the performance of the BERT architecture. RoBERTa replaces the WordPiece tokenizer used in the original BERT with Byte-Pair Encoding (BPE) for generating the token vocabulary. It eliminates the NSP task from pretraining and modifies the MLM task by incorporating dynamic masking. Additionally, RoBERTa leverages a significantly larger dataset, longer training duration, larger batch sizes, and an optimized learning rate schedule to achieve superior performance.

#### Closed-source models

Compared to open-source models, closed-source models generally provide more powerful solutions for generating embeddings. This is because they are often trained with large-scale, high-quality proprietary datasets and leverage advanced architectures and training techniques that are not publicly accessible.

The superiority of those models compared to older generation open sourced models, such as BERT and RoBERTa, is evident in the text retrieval benchmark leaderboard, MTEB ^7,17^. In this study, we chose the encoder-only models from Cohere, Google, and OpenAI (**Supplementary Table 2**). However, despite those models being publicly available for deployment through web interfaces, the proprietary nature of those models makes some parameters less transparent, hence difficult to compare. Therefore, we choose models from those three companies based on the versioning provided (**Supplementary Table 2**).

## Comparing Encoded Spaces

The distances between encoded texts are defined by the cosine similarities of the embedded values. For each pair of the texts, the correlations between two vectors of the embedded values are calculated. The pairwise distances of the input texts inform us about the relative positions of each item in the embedded space. By comparing the distances in the text embeddings (cosine similarities) and the distances in the biological embeddings (genetic correlations), we can evaluate how well those two embeddings align with each other. We used linear mixed effects meta-analysis to evaluate the degree of the alignment between two types of the embeddings.

## Acknowledgement

This work was partly funded by The William K. Warren Foundation, the National Institute of General Medical Sciences Center (Grant 2 P20GM121312, MPP, RK, KLF), the National Institute on Drug Abuse (U01DA050989, MPP), and the National Institute of Mental Health (R01MH122688, R01MH128959, CCF). Dr. Paulus advises Spring Care, Inc., receives royalties from an article on methamphetamine in UpToDate, and has a compensated consulting agreement with Boehringer Ingelheim International GmbH.

---

**Supplementary Table 1.**

| Model Families | Name | Context window size | Embedding dimension | MTEB* average score |
| --- | --- | --- | --- | --- |
| Open-source<sup>‡</sup> | BERT | 512 | base: 768<br>large: 1024 | BERT<sub>base</sub>: 38.33<br>other: NA |
|  | RoBERTa |  |  | NA |
| Cohere<sup>§</sup> | v2 light | 512 | 1024 | NA |
|  | v2 english |  | 4096 | NA |
|  | v3.0 light |  | 384 | 62.0 |
|  | v3.0 large |  | 1024 | 64.5 |
| Google<sup>†</sup> | Gemini | 2048 | 768 | 66.3<sup>†</sup> |
| OpenAI<sup>□</sup> | ada v2 | 8191 | 1536 | 61.0 |
|  | 3-small |  | 1536 | 62.3 |
|  | 3-large |  | 3072 | 64.6 |

\* Massive Text Embedding Benchmark (MTEB): a massive benchmark for measuring the performance of text embedding models on diverse embedding tasks, including 56 datasets across 8 tasks <sup>7</sup>.
<sup>‡</sup> Scores for BERT<sub>large</sub>, RoBERTa<sub>base</sub> or RoBERTa<sub>large</sub> are not published on the MTEB Leaderboard <sup>18</sup>.
<sup>§</sup> MTEB scores for embed-english-light-v2.0 and embed-english-v2.0 are not released by Cohere.
<sup>†</sup> The specific MTEB score for Google’s text-embedding-004 model is not publicly available. However, the Gecko embedding models <sup>19</sup>, which are also part of Google’s text embedding family, have reported MTEB average scores of 64.4 for the gecko-1b-256 model and 66.3 for the gecko-1b-768 model.
□ Text-embedding-ada-002 and text-embedding-3-small/large are released in 2022 and 2024 separately

**Supplementary Table 2.**
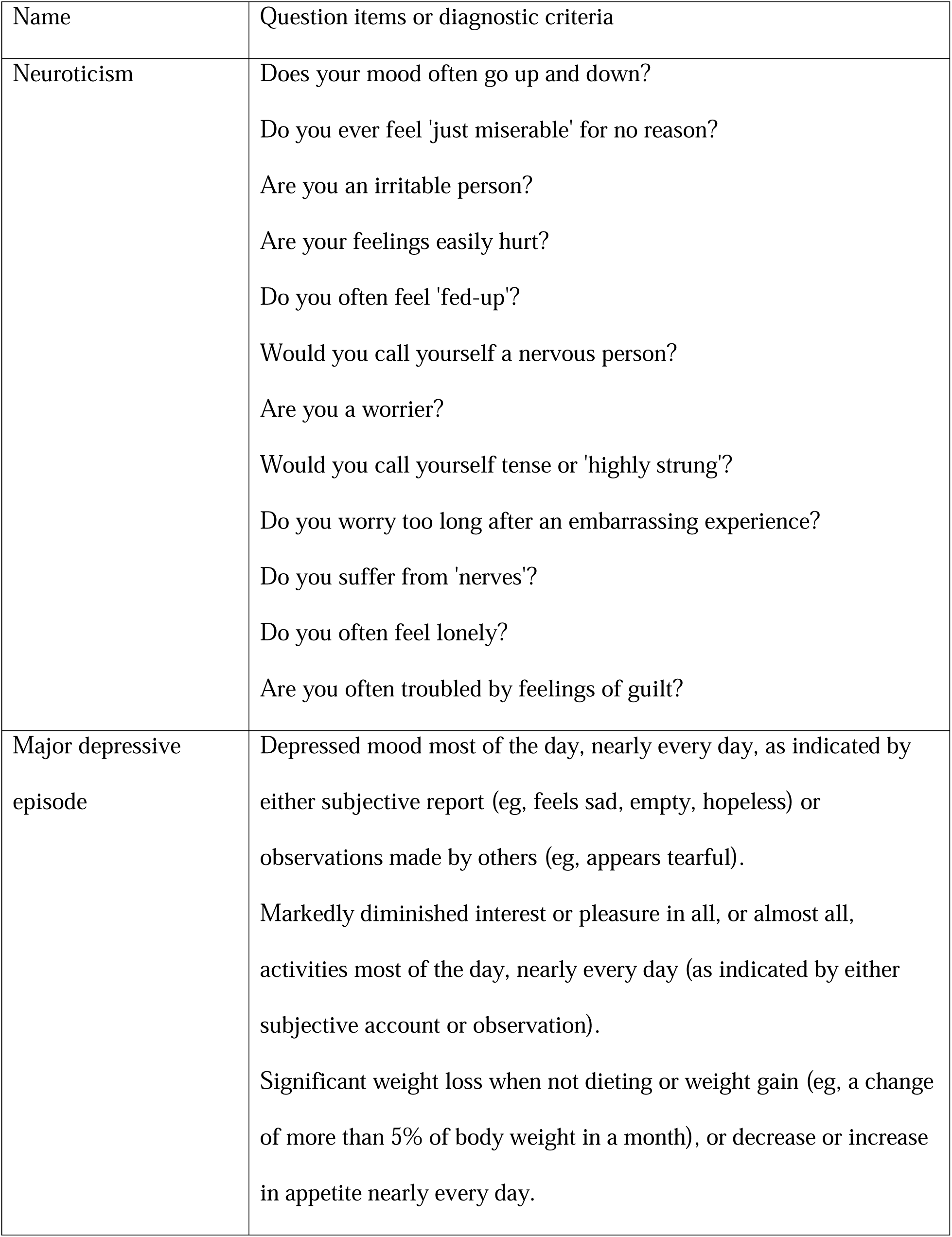
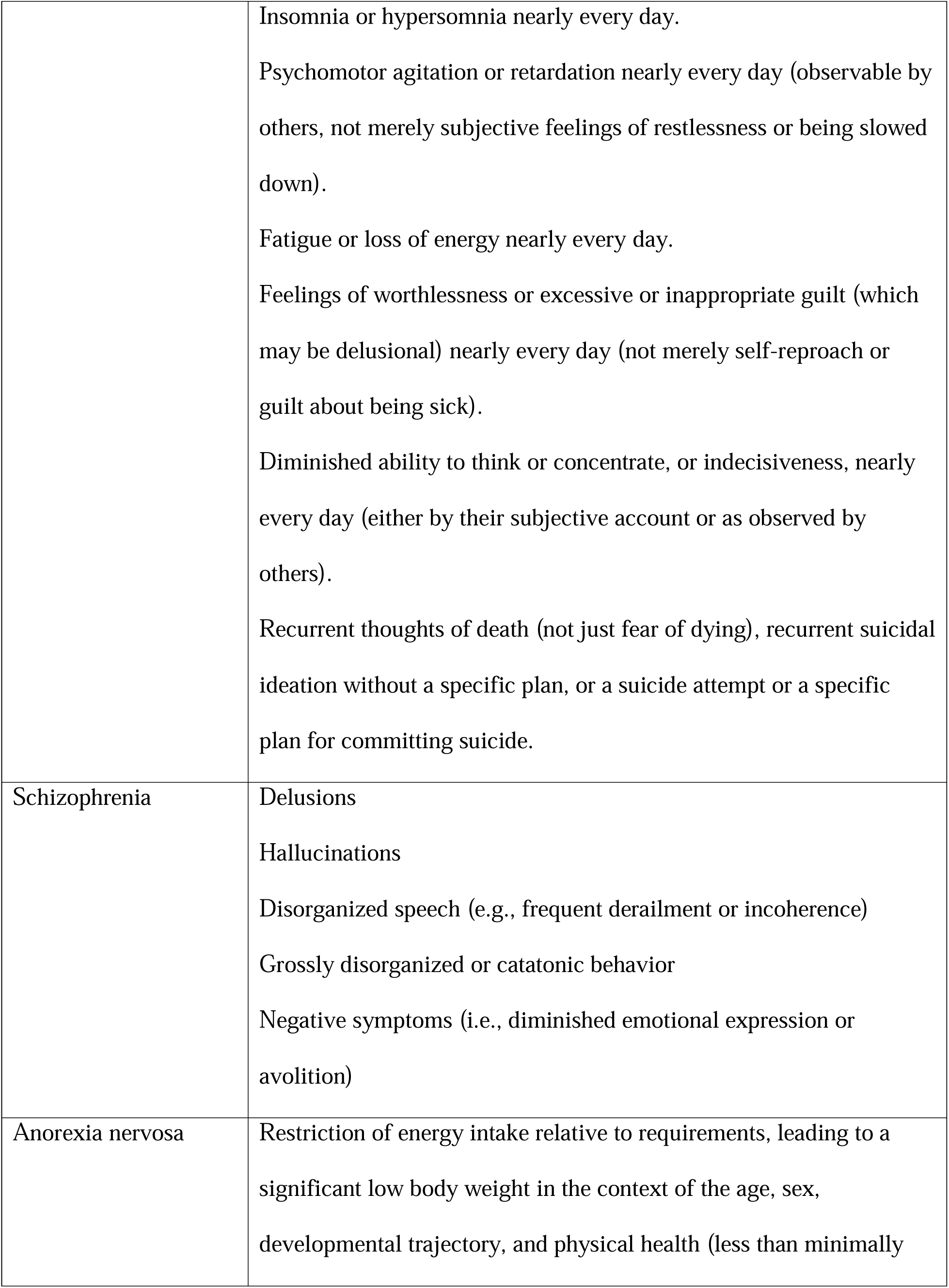
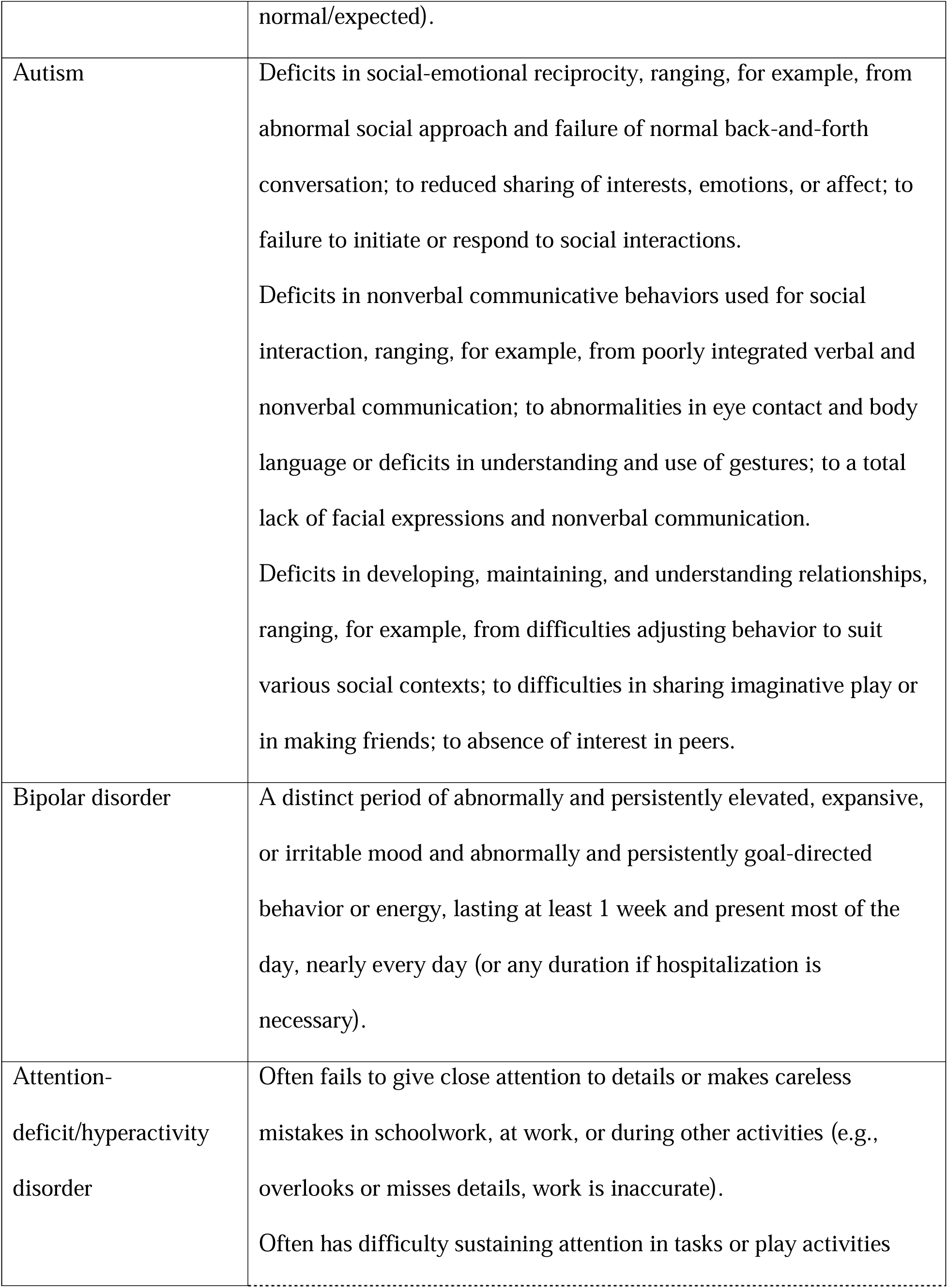
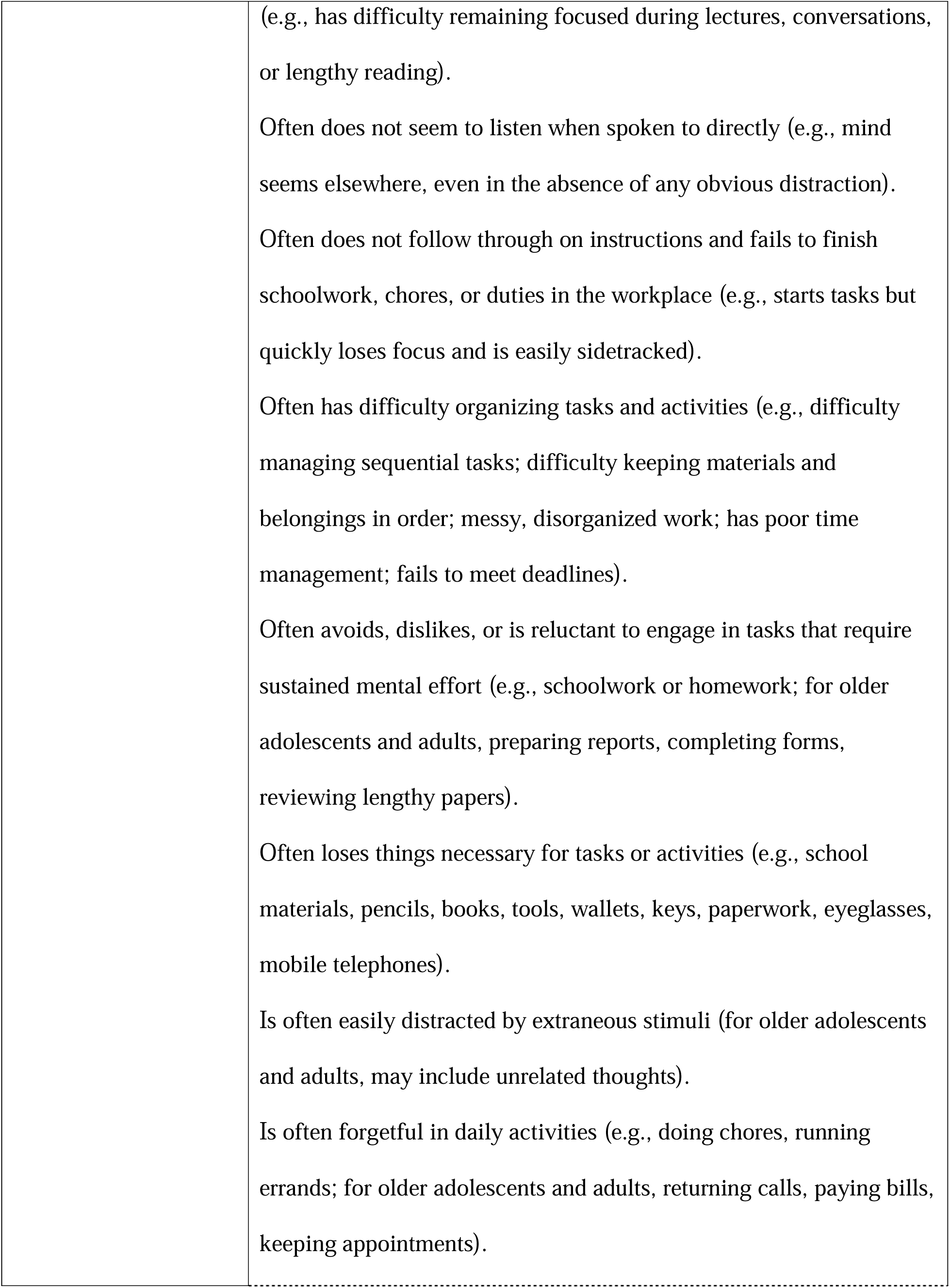
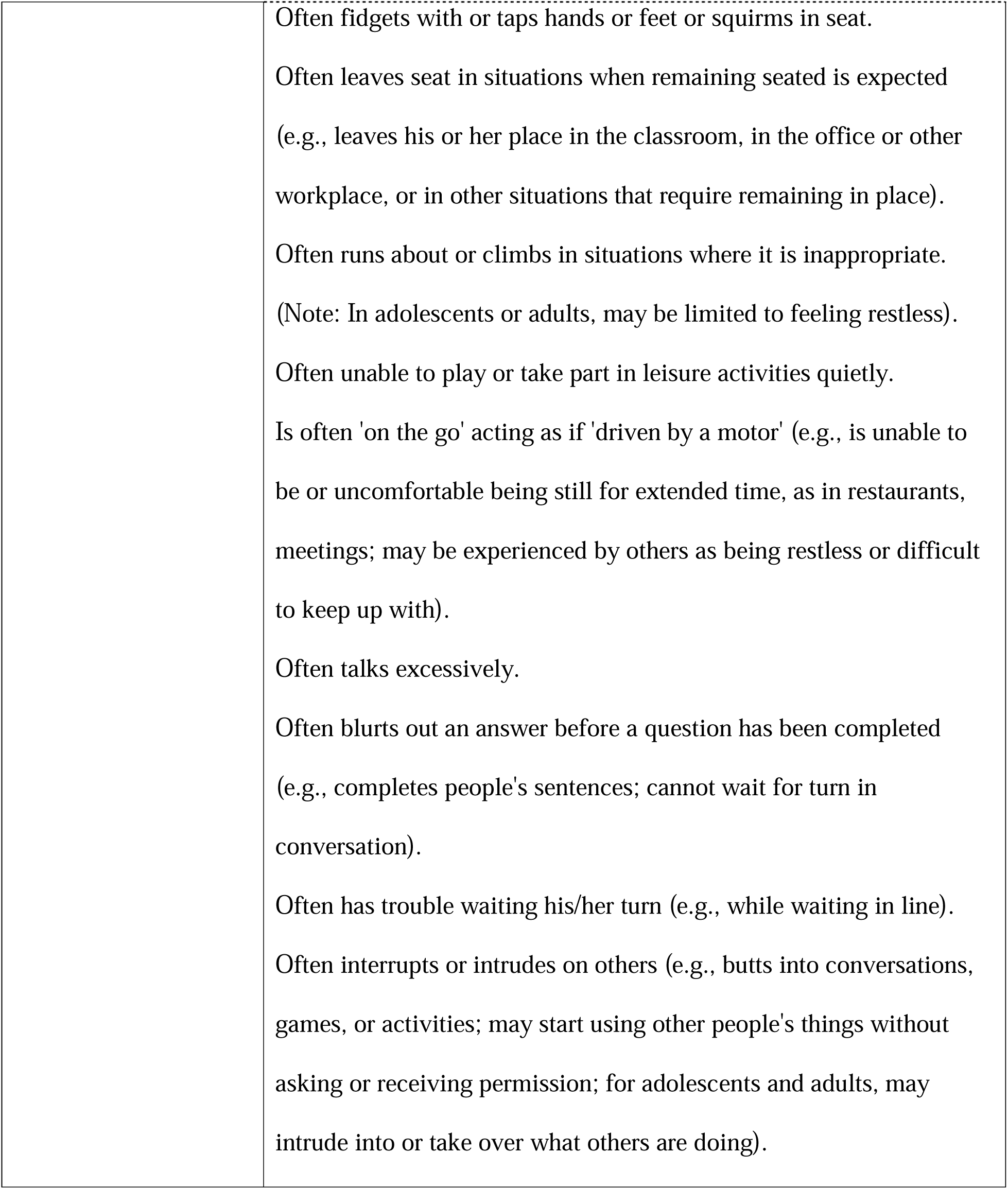

